# Containing the Spread of Infectious Disease on College Campuses

**DOI:** 10.1101/2020.07.31.20166348

**Authors:** Mirai Shah, Gabrielle Ferra, Susan Fitzgerald, Paul J. Barreira, Pardis C. Sabeti, Andres Colubri

## Abstract

College campuses are highly vulnerable to infectious disease outbreaks, and there is a pressing need to develop better strategies to mitigate their size and duration, particularly as educational institutions around the world reopen to in-person instruction during the COVID-19 pandemic. Towards addressing this need, we applied a stochastic compartmental model to quantify the impact of university-level responses to past mumps outbreaks in college campuses and used it to determine which control interventions are most effective. Mumps is a very relevant disease in such settings, given its airborne mode of transmission, high infectivity, and recurrence of outbreaks despite availability of a vaccine. Our model allows for stochastic variation in small populations, missing or unobserved case data, and changes in disease transmission rates post-intervention. We tested the model and assessed various interventions using data from the 2014 and 2016 mumps outbreaks at Ohio State University and Harvard University, respectively. Our results suggest that in order to decrease infectious disease incidence on their campuses, universities should apply diagnostic protocols that address false negatives from molecular tests, stricter quarantine policies, and effective awareness campaigns among their students and staff. However, one needs to be careful about the assumptions implicit in the model to ensure that the estimated parameters have a reasonable interpretation. This modeling approach could be applied to data from other outbreaks in college campuses and similar small-population settings.

## 1 Introduction

College campuses provide ideal breeding grounds for infectious disease. Students live in close quarters, pack into lecture halls, share food and drinks in the dining areas, and engage in intimate contact. Outbreaks in these settings can spread very quickly. Indeed, a meningitis outbreak took place at Princeton University in March 2014, eventually claiming the life of one student. The Centers for Disease Control and Prevention (CDC) reported the attack rate of the disease on Princeton’s campus to be 134 per 100,000 students – 1,400 times greater than the national average (1). Recent COVID-19 spread in educational settings (2) forced school closures around the world (3), and motivated the design and implementation of plans for safe reopening (4, 5).

A recent string of outbreaks on college campuses involves mumps, once a common childhood viral disease. After introduction of the measles-mumps-rubella (MMR) vaccine in 1977 and the two-dose MMR vaccination program in 1989, the number of mumps cases in the US plummeted by 2005. But, despite a vaccinated population, there has been a recent resurgence of mumps, with a steep jump from 229 cases in 2012 to 5833 cases in 2016 (6). Although a typically mild disease in children, up to 10% of mumps infections acquired after puberty can cause severe complications, including orchitis, meningitis, and deafness. Furthermore, a majority of recent mumps cases have occurred in young adults who had received the recommended two MMR doses. This suggests that vaccine-derived immunity wanes over time, unlike natural immunity – protection acquired from contracting the disease – which is permanent. Lewnard and Grad estimate that 33.8% of young adults (ages 20 to 24) were susceptible to mumps in 1990, in contrast to the 52.8% susceptible in 2006, as vaccinations have replaced contraction as the source of immunity (7). The temporary immunity from vaccines strengthens the argument for strict containment as a critical line of defense amidst an outbreak. In the case of COVID-19, even with the availability of several vaccines (8), the challenges associated with their wide and quick distribution (9), the substantial asymptomatic and pre-symptomatic transmission of the disease (10), and the possibility of new viral strains with higher transmissibility (11) provide further support for such approaches.

The spread of mumps at Harvard University in 2016, and extensive public health measures and documentation, presents an opportunity to closely examine an outbreak on a college campus. Between January 1 and August 31, 2016, 210 confirmed mumps cases were identified in the Greater Boston area, with most detected at Harvard University. Mumps is a highly contagious disease with the potential to travel quickly and pervasively on a crowded college campus. Some of the most notable mumps outbreaks on college campuses occurred in Iowa (12), Indiana (13), and Ohio (14). But, whereas mumps spread rapidly at Ohio State University (OSU) in 2014 and the University of Iowa in 2006 and 2016, Harvard employed a number of interventions that may have helped mitigate spread of the disease and contain it over just a few months (15). The possibility of distinct viral strains resulting in different outbreak dynamics between schools can be safely dismissed, as it was shown by application of genetic epidemiology methods (16) that all mumps outbreaks in the US since at least 2006 have been likely caused by the same mumps lineage, mumps virus genotype G.

The successful containment at Harvard motivates us to explore varied intervention strategies, given the relative costs of prevention. Even if the use of a booster MMR vaccination is proven theoretically to reduce infection and thus potentially prevent outbreaks (7, 12), it is unlikely that universities with limited resources will proactively invest in a third dose. A rough cost analysis conducted by Harvard University Health Services (HUHS) showed that, while the total mumps care expenses for Harvard was approximately $75,000, the cost of providing a third MMR dose to every member of the Harvard community (at $83 per vaccination) was $1.7 million (17). Therefore, at least in the short term, a third MMR dose cannot be the only answer to handling mumps outbreaks; we must consider more immediate solutions and interventions.

In order to understand the effectiveness of interventions aimed at containing mumps outbreaks on a college campus, we constructed an epidemiological model to simulate the dynamics of mumps on such a population and quantify the impact of various interventions. This modeling can be challenging because the number of susceptible students in a college is small compared with population-wide studies, parameters characterizing the interventions are not known, and data is partially observed. Deterministic models are only appropriate when the populations of the compartments are sufficiently large (18). We adopt a modified stochastic susceptible-exposed-infectious-recovered (SEIR) model presented to addresses these issues. We developed this model within the framework of a Partially Observed Markov Process (POMP), which has been applied to introduce structural stochasticity into epidemic models (19). The stochastic nature of the model allows for variability from elements that are not explicitly included, such as class schedules and campus layouts. This model also allows to easily quantify time-varying interventions after fitting the parameters to the observed data, by running simulations under alternative scenarios.

We fit model parameters on case data for Harvard’s 2016 mumps outbreak provided by the Massachusetts Department of Public Health (MDPH). We compared it to data from OSU, one of the few universities that had extensive publicly available data through the CDC.

In applying our model, we found that each of the interventions employed by HUHS -- email awareness campaigns, more aggressive diagnoses where clinical symptoms alone were enough to result in quarantine, and strict isolation of suspected cases -- were crucial in reducing the size and duration of the outbreak. In particular, Harvard’s policies drastically increased the reporting rate of infection and shortened the time a person remains infectious in a susceptible population, relative to the baseline. As a result, one mumps case at Harvard infected less than two susceptible individuals on average, and much less once aggressive diagnosis was in place, compared to cases at non-residential schools like OSU, in which one mumps case infected an average of six susceptible individuals. However, the OSU data suggests that self-isolation could be effective, if adopted rigorously by students. The conclusions from this paper could guide future responses to infectious disease outbreaks on college campuses. Without effective measures in place, highly transmissible diseases like mumps, meningitis, and now COVID-19, spread in these environments at much faster rates than in the overall population and can lead to serious health complications. Simple interventions that ensure most cases are detected, treated, and separated from susceptible individuals make a significant difference.

## 2. Materials and Methods

### 2.1 Harvard mumps outbreak

#### 2.1.1 Data

The mumps outbreak at Harvard began in February 2016, when six students reported onset of parotitis to HUHS. For the next three months, the number of cases continued to rise, until finally plateauing in late May and early June. There were two waves of the outbreak – one occurring in the month of March and a larger one occurring in mid-April – totaling 189 confirmed and probable cases (Figure 1). Confirmed cases are those with a positive laboratory test for mumps virus. Probable cases are those who either tested positive for the anti-mumps IgM antibody or had an epidemiologic linkage to another probable or confirmed case (20, 21). The majority of these cases received the recommended two doses of MMR (22).

**Figure 1:**
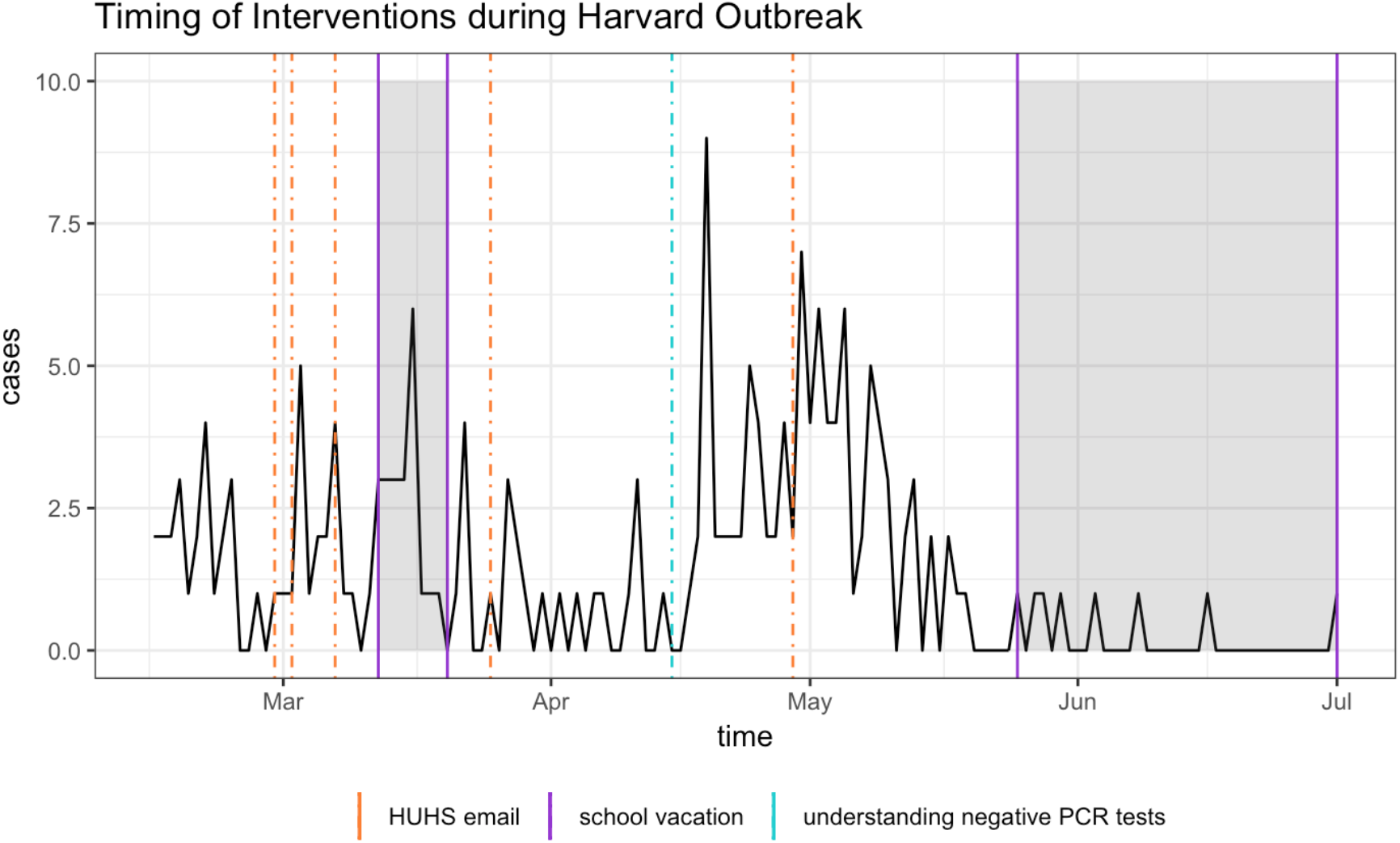
The daily number of new mumps cases (probable or confirmed) at Harvard and the timeline of school vacations and control interventions employed by HUHS between February and June 2016. Both probable and confirmed cases display clinical symptoms of mumps, but only confirmed cases have a positive PCR result. HUHS sent multiple emails over the course of the outbreak, raising awareness about the spread of mumps. Additionally, in mid-April, HUHS began more carefully diagnosing mumps, rather than automatically ruling out those with negative PCR tests. The isolation policy is not shown because it occurred continuously throughout the entire outbreak.

We use data provided by MDPH, which documented every mumps case between 2015 and 2017 at schools across Massachusetts (23). This data includes demographics of the patient (gender, age, county, and institution), symptoms and vaccination status, date they reported their symptoms and the date of symptom onset, and lag time between the date of symptom onset and admission to a medical clinic.

#### 2.1.2 Interventions

Harvard University employed three main interventions: (i) an email awareness campaign, (ii) more aggressive diagnoses, and (iii) strict isolation of infectious persons.

First, between February and May 2016, HUHS sent six different emails to Harvard students, employees, and colleagues with information on the gravity of the outbreak, recommendations on how to prevent transmission, and instructions on how to identify mumps. This raised awareness throughout the campus. Particularly at the peak of the outbreak, roommates, resident deans, and athletic coaches all played essential roles in reporting potential cases of mumps, so that few cases likely went undetected and untreated by HUHS (20, 21).

Second, Harvard acted vigorously to treat and isolate anyone suspected of mumps throughout the outbreak. Initially, due to the disease’s non-specific symptoms and less extreme manifestation in vaccinated people, HUHS used positive mumps PCR tests as a necessary ground for diagnosis. Later, on recommendation from the MDPH, HUHS stopped automatically ruling out those with negative PCR results, given that false negatives were quite frequent in vaccinated individuals and that some individuals reported their infection to the clinic belatedly. Isolation or detection of the mumps virus is challenging because the of its transient replication and coincident presence of antibodies (24). In outbreaks among two-dose vaccine recipients, mumps virus was only detected in samples from approximately 30-35% of case patients if the samples were collected within the first three days following onset of parotitis (25). Anyone who entered HUHS displaying clinical symptoms of mumps was now deemed infected and infectious. This change in the diagnosis protocol took place on April 15, 2016, day 61 of the outbreak (21).

Third and perhaps most notably, Harvard isolated most confirmed or probable cases of mumps. While many universities simply suggest self-isolation in one’s room or dormitory (which leaves roommates and friends highly susceptible to the disease), Harvard removed anyone with clinical symptoms of mumps from the population. Of the 230 total cases at Harvard between February 2016 and November 2017, 96 were isolated in alternate housing on campus, while 110 were isolated off-site. Although a person remains infectious with mumps for five days, Harvard isolated patients for six days for additional measure (20).

Harvard also used a variety of smaller techniques to contain the disease. For instance, water fountains with a weak upward flow were repaired in late March when it became apparent that students were directly touching the fountain with their water bottles or mouths (21). In this study, we only considered the first three larger-scale interventions in our models. Figure 1 shows a timeline of the interventions as well as periods when the population was fluctuating (such as during spring and summer break). Around two weeks after HUHS improved its criteria for diagnosis in mid-April, there was a steep decline in the number of new cases. These interventions were possible thanks to the ample resources that Harvard has at its disposal, which may not be available at other universities. Nevertheless, this situation makes Harvard an ideal testing ground for interventions that could not be deployed elsewhere, at least without solid proof of their efficacy. Thus, we quantify the effects of the three main interventions (awareness campaign, aggressive diagnoses, and strict isolation of suspected cases) further in the modeling section of this paper.

### 2.2 Ohio State University mumps outbreak

#### 2.2.1 Data on the outbreak

In 2014, a large outbreak of mumps occurred in central Ohio, with the majority of cases linked to OSU in Columbus. The outbreak began in February 2014 and peaked in early April with 96 cases in one week. By summer and early fall, the number of cases had dramatically dropped and stabilized (14). We therefore restrict our analysis of the outbreak to the time between Week 1 and Week 40 of 2014, in which there were a total of 528 cases (Figure 2). We obtained this data from CDC’s *Morbidity and Mortality Weekly Report* (26). One drawback of the data is that the cases are reported weekly, making our analysis and parameter estimations less precise. Furthermore, we cannot guarantee that all the cases in this dataset are linked to the university itself, but we know from news reports that most cases in Ohio occurred on campus during the first half of 2014 (14). The proximity in time to the Harvard outbreak and the differences in response detailed below make this a good dataset to compare to.

**Figure 2:**
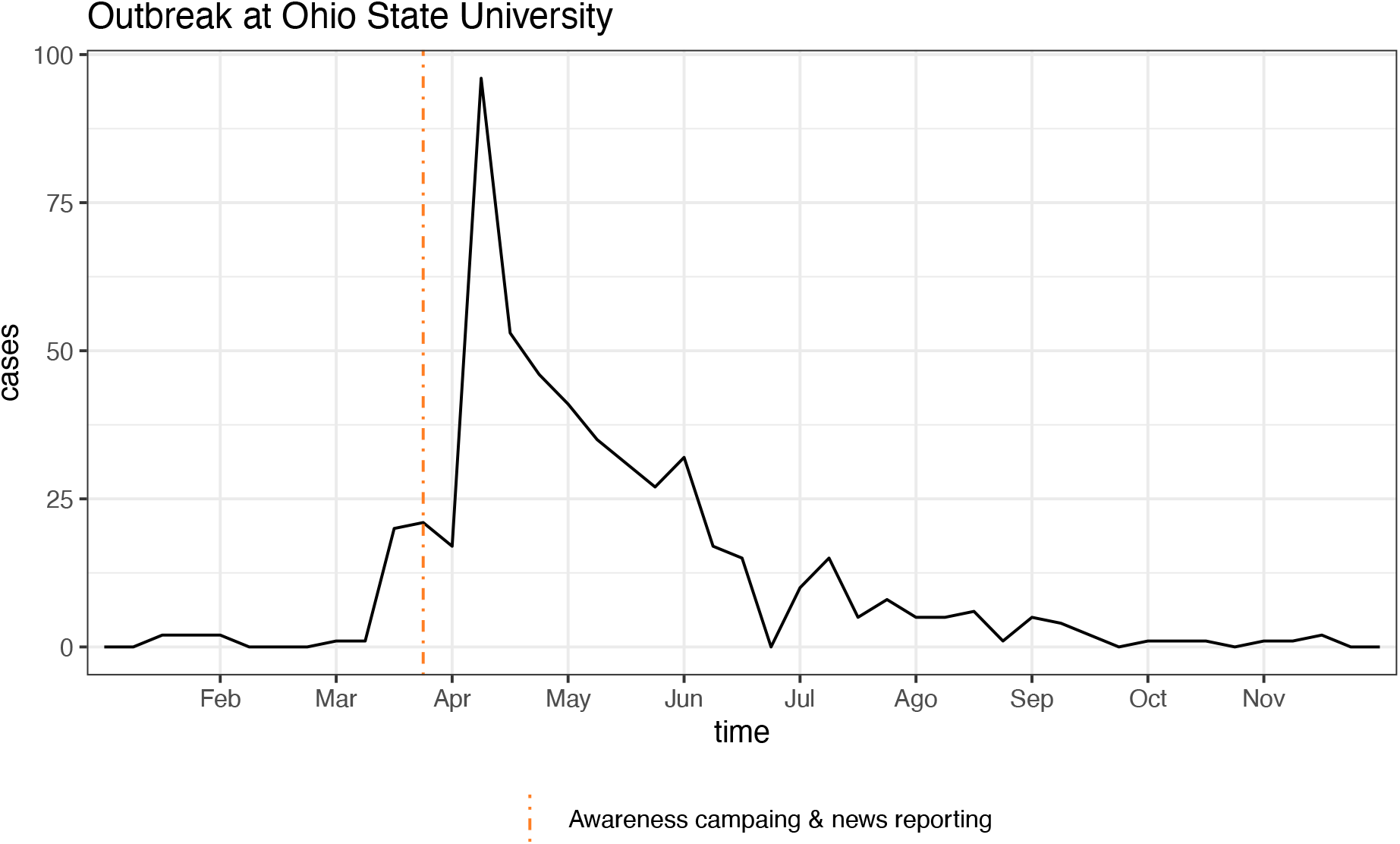
Number of weekly mumps cases in Ohio (particularly Ohio State University) between January and November 2014. There were 528 cases during this time period, with most occurring between Match and July. The dotted line in the last week of March indicates the intervention consisting in awareness campaign by OSU, as well as local and national news reports about the outbreak.

#### 2.2.2 Characteristics of the response

We were unable to acquire data directly from OSU, and thus the exact timeline and range of interventions administered over this period are not known. We learned through online searches that advisories were published by the university, notifying students of the issue and how to prevent its spread. One notice published by OSU’s medical center reads: “Stay at home for five days after symptoms (salivary gland swelling) begins (required by Ohio law OAC 3701-3-13, (P)); avoid school, work, social gatherings, and other public settings” (27). These advisories were distributed since March 2014 (28), and local news outlets also started reporting the outbreak earlier in the month (29). It appears, however, that like most affected universities, OSU did not formally isolate infectious persons.

### 2.3 Epidemiological POMP model

The epidemiology of mumps can be captured by a Susceptible-Exposed-Infected-Removed (SEIR) compartmental model: after exposure, individuals go through a latent non-infectious period, followed by an infectious phase (30). Infectious individuals are removed from the transmission process either by recovery or isolation, after which they become immune. Compartmental models simplify the mathematical modeling of infectious diseases; however, they assume access to fully observed disease data. In reality, not all mumps cases are reported, and latent mumps carriers exhibit no symptoms at all. In order to address this issue, our approach integrates a standard SEIR model with a Partially Observed Markov Process (POMP) model (31). This allows us to combine the simplicity of compartmental models with a probabilistic framework for the underlying dynamics and the observed data. POMP models require the specification of a process model that describes stochastic transitions between the (unobserved) states of the system (in this case, the SEIR compartments), and a measurement model where the distribution of observed data (e.g.: confirmed cases) is expressed as a function of the unobserved states. The stochasticity introduced in the SEIR dynamics makes our model better suited to describe small populations, such as college campuses, where random fluctuations can be significant in relation to the size of the population. We describe the process and measurement models below.

#### 2.3.1 Process model

The process model, defined as a stochastic SEIR model, provides the change in true incidence of mumps at every time point. We add parameters that induce random fluctuations into the population and change the compartments’ rates of transfer in response to interventions. We do this by using probabilistic densities for the transition of state variables. Moreover, although disease dynamics are technically a continuous Markov process, this is computationally complex and inefficient to model, and so we make discretized approximations by updating the state variables after a time step, *δ*. Due to the varying granularity of the observed data (daily and weekly), we used two different time steps: *δ*_*H*_=2.4 *hours* for Harvard and *δ*_0_=12 *hours* for OSU. The system of discretized equations is shown in Equation 1, where *B*(*t*) is the number of susceptible individuals who become exposed to mumps, *C*(*t*) is the number of newly infectious cases, and *D*(*t*) is the number of cases that are removed from the population:

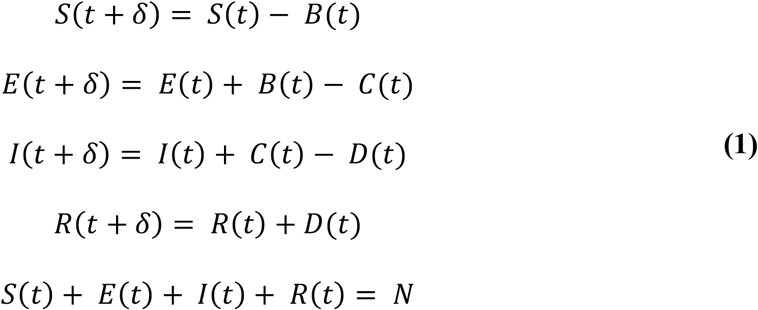

Equation 1 describes how the sizes of the four compartments (susceptible, exposed, infectious, and removed) change between (*t, t*+*δ*). The model further assumes that the population size *N* remains constant at every time point. We added inherent randomness to our model by setting *B*(*t*), *C*(*t*), and *D*(*t*) as binomials. If we assume that the length of time an individual spends in a compartment is exponentially distributed with some compartment-specific rate *x*(*t*), then the probability of remaining in that compartment for an additional day is *exp*(−*x*(*t*)) and the probability of leaving that compartment is 1 − *exp* (−*x*(*t*)):

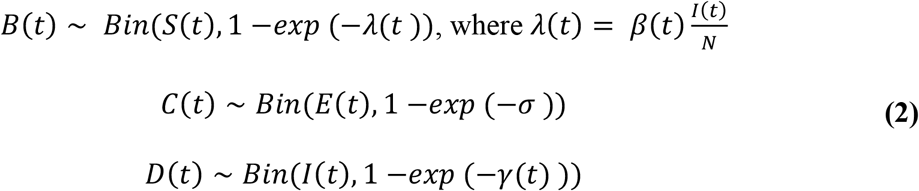

The force of infection, *λ*(*t*), is the transition rate between the susceptible and exposed classes at time t and can be expressed as 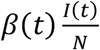, where *β*(*t*) represents the transmission rate of the disease. The removal rate between the infectious and removed compartments at time t is given by *γ*(*t*), and transition rate between the exposed and infectious classes is *σ*. Therefore, *γ*(*t*)^−1^ represents the mean length of time a person is infectious before being removed from the population (either because of intervention efforts or natural recovery), while *σ*^−1^ represents the mean length of time a person stays in the latent stage. With this notation, we are implicitly assuming that the transmission and removal rates could change over time due to interventions or changes in behavior, while the duration of the latent stage is constant and determined by the physiopathology of the disease. We will justify these assumptions for Harvard and OSU next, as well as provide explicit formulas for *β*(*t*) and *γ*(*t*).

Leaving aside the unlikely possibility of change in pathogen’s infectivity, the transmission rate *β*(*t*) essentially depends on the frequency of exposure events. In the case of Harvard, its nature as a residential campus would lead to significant decreases in student population, and therefore exposures, during school vacations. Exposure at OSU, a non-residential campus, is arguably less affected by vacation breaks. Another potential cause for reduction in exposures is awareness campaigns resulting in the adoption of preventive behaviors by students. Both Harvard and OSU adopted such campaigns, in the former, implemented as emails regularly sent out by HUHS recommending personal hygiene and testing in case of symptoms compatible with mumps; in the latter, in the form of advisories posted around campus and online, advising self-isolation to those students who presented symptoms. Furthermore, due to the scale of the mumps outbreak in Ohio, it received local and national news coverage, particularly in connection with OSU. Anecdotal evidence (i.e.: conversation with students) and, most importantly, the fact that HUHS emails were throughout the outbreak, make us conclude that emails were not particularly effective. On the other hand, news coverage in the case of OSU could have led to additional awareness by students and encouraged some to self-isolate. We argue that self-isolation results in lowering of transmission rate, not shortening of the removal time, because it is not perfect quarantine and people can still interact and become exposed, albeit at a lower frequency. Based on these known facts and our interpretation of them, we propose the following transmission rate *β*_*H*_(*t*) for the Harvard model:

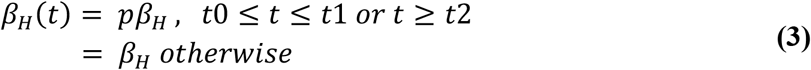

Here, *t0* and *t1* represent the starting and ending dates for the spring break (March 12-20 2016), and *t2* the beginning of the summer recess (May 26 2016). The constant *β*_*H*_ is the baseline transmission rate during normal class term, and the parameter *p* is a number between 0 and 1 that accounts for the reduction of student population on campus during the school vacation. In the case of OSU, we propose:

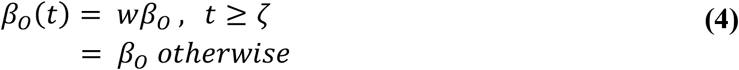

In this equation, *β*_0_ the baseline transmission rate, *w* is a constant lower than 1, and *ζ* the time when students began to self-quarantine. Based on publication of public health advisories and local news, we set this time as the last week of March 2014 (week 12). Since Harvard’s quarantine was in effect through the entirety of the outbreak, we did not incorporate a similar *w* coefficient to the corresponding *β*_*H*_(*t*) equation for Harvard.

The removal rate *γ*(*t*) can also be affected by interventions and personal behaviors. We know that HUHS diagnosis protocol changed on day 61 of the outbreak at Harvard, resulting in a shorter average removal time since clinical presentation of symptoms alone was enough to result in strict isolation of suspected cases. Thus, we propose the following *γ*_*H*_(*t*) for Harvard:

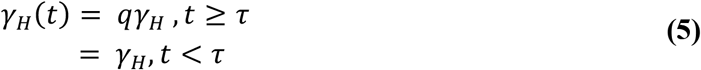

Here, *q* is a constant greater than 1 and *τ* is the date when the new criteria was implemented (April 15, 2014). The constant *γ*_*H*_ is the baseline removal rate reflecting the impact of the original diagnosis protocol. In the OSU model, on the other hand, we assume a constant recovery rate *γ* equal to the population average for mumps, since infected individuals self-isolate at home. This would not result in a strict quarantine but in a reduced contact rate with susceptible individuals, which is already modeled by a lower transmission rate in equation (4).

Finally, it is necessary to estimate the basic reproduction number, *R*0, which equals the expected number of secondary cases produced by an infectious person in a completely susceptible population (30). *R*0 measures the initial growth rate of an outbreak and so, if it is less than 1, then the infection will die out and there will be no epidemic. For our stochastic SEIR model, this constant can be expressed as 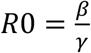. Meanwhile, the time-dependent effective reproduction number is defined as 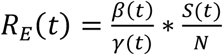, but because *S*(*t*) ≈ *N*, we can simplify this expression to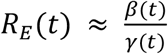. Both the basic and effective reproduction numbers allow us to understand the strength of an outbreak.

#### 2.3.2 Measurement Model

Although it is impossible to directly record the number of people that are susceptible, exposed, infectious, and removed directly, the MDPH and CDC data tells us the number of observed cases per day. The mean number of observed cases per day is the true number of cases multiplied by the reporting rate *ρ* (*ρ* < 1). However, rather than simply denoting the observed number of cases as a binomial distribution, we account for greater variability in the measurements than a binomial distribution expects, since college populations are “small” (comparted to cities and larger administrative units) and more affected by random fluctuations (32). Thus, the number of observed cases, *y*_*t*_, given the number of true cases, *C*(*t*), can be best modelled by an overdispersed binomial distribution defined as a discretized Normal random variable:

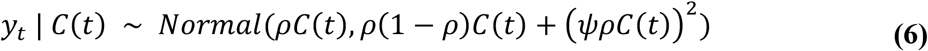

The parameter *ψ* handles the increased variability in a small population. If *ψ*=0, the variance in our measurement model simplifies to the variance for a binomial distribution.

#### 2.2.3 Final POMP Model

The process and measurement models define our final POMP model. For each time point, the process model generates the number of new cases based on binomially distributed counts. The measurement model then estimates the observed number of cases based on the true number of cases and reporting rate. The free parameters in our POMP models for Harvard and OSU that need to be estimated from the data are the following: (i) *β*_*H*_ and *β*_0_, baseline transmission rates, (ii) *p* and *w*, decrease in transmission rate at Harvard and OSU due to vacation and self-isolation, respectively, (iii) *γ*_*H*_ baseline removal rate at Harvard (iv) *q*, increase in removal rate due to the updated HUHS diagnosis protocol, (v) *ρ*_*H*_ and *ρ*_0_, case reporting rates, (vi) *ψ*_*H*_ and *ψ*_0_, overdispersion coefficient representing additional variability in the populations.

### 2.3 Fixed parameters

In addition to the free parameters to be estimated from the observed case data, our models also include a number of fixed parameters, shown in Table 1, whose values can be inferred directly from previous knowledge or available information. As mentioned earlier, we chose *τ*=61 days and *ζ*=12 weeks because those points in time at Harvard and OSU correspond to the introduction of the interventions that we hypothesized to be impactful in the dynamics of the respective outbreaks. Dates *t0, t1*, and *t2* for the spring and summer vacations at Harvard are available online (33). We set the rate between the exposed and infectious classes and the recovery rate to 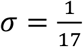 and 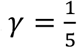, respectively, since the average latent period and recovery time for mumps are known to be *σ*^−1^=17 days and *γ*^−1^=5 days (7). Finally, we set the effective population size at Harvard *N*_*H*_=20,000 × 0.53=10,600 people based on records of Harvard’s enrollment and employment, and Lewnard and Grad estimation of susceptibility to mumps among college-age adults due to immunity waning (7). Similarly, we use an effective population for OSU given by *N*_0_=60,000 × 0.53=31,800, leveraging the total enrollment for the 2013-2014 academic year reported in OSU’s statistics website (34).

**Table 1:** List of fixed parameters used in mumps transmission model for Harvard and OSU

| *Symbol* | *Description* | *Value* | *Units* | *Source* |
| --- | --- | --- | --- | --- |
| $\tau$ | Date of intervention at Harvard | 61 | day | Harvard records on interventions (21) |
| $t_0, t_1, t_2$ | Vacation dates at Harvard, 2015-2016 academic year | 26, 34, 100 | day | Harvard archived academic calendar (33) |
| $\zeta$ | Date of intervention at OSU | 12 | week | |
| $\sigma^{-1}$ | Duration of mumps latent period | 17 | day | Lewnard and Grad (7) |
| $\gamma^{-1}$ | Duration of mumps recovery period | 5 | day | Lewnard and Grad (7) |
| $N_H$ | Effective population at Harvard | 10,600 | — | Harvard records on population size (22) and mumps susceptibility among college-aged individuals (7) |
| $N_O$ | Effective population at OSU | 31,800 | — | OSU's statistical summary (34) and mumps susceptibility among college-aged individuals (7) |

### 2.4 Maximum likelihood estimation of free parameters

In order to obtain estimates of the free parameters in our models, we pick the parameter values that maximize the log likelihood of the observed data given each model. Within the POMP framework, we can perform fast maximum likelihood estimation (MLE) via Sequential Monte Carlo (SMC) techniques (31). SMC allows us to calculate the likelihood of the data more efficiently by applying the Markov property to generate paths in parameter space that sample the likelihood surface. We performed 100 searches from random parameter guesses, each converging to a unique value, and we then took the maximum over the 100 runs the final point estimates. We did this using the pomp package version 3.3 (35) for the R statistical software version 4.0.5 (36). In order to calculate the confidence intervals for each parameter, we applied the Monte Carlo-adjusted profile (MCAP) algorithm (37).

### 2.5 Intervention analysis

Finally, we performed an analysis of the parameters *q* and *w*, which respectively quantify the effect of what we consider to be the defining intervention at Harvard (aggressive diagnosis) occurring around day 61 of the outbreak, and the self-isolation awareness campaign at OSU during March 2014. This could allow us to understand to what extent these interventions made a difference on the trajectory of the outbreak. First, we compared the scenario with the interventions versus a scenario without the interventions. Controlling for all other parameters, we run two sets of simulations at the MLEs, with 200 simulations each. The first set of simulations fixed *q* and *w* at the value obtained from MLE, while the second set of simulations set *q* and *w* to 1, assuming that no interventions occurred around day 61 at Harvard and by week 12 at OSU. We then compared the cumulative number of cases over time for these two sets of simulations, generating a 95% percentile range from all the simulations in each set. Second, we used this method to determine if administering the interventions earlier could have lowered the number of cases. For Harvard, we let the day of the intervention take on values between 1 and 60. Subsequently, we ran simulations for each of these 60 cases, pulled the final outbreak size from the median simulation, and calculated the reduction in outbreak size. We applied the same procedure for OSU, in this case varying the day of intervention between 1 and 11 and calculating the corresponding final outbreak sizes.

## 3. Results

### 3.1 Optimal Parameters of Harvard and OSU Outbreaks

The MLEs of the parameters provide insight into the key characteristics of Harvard’s and OSU’s outbreak. In general, we observe very good agreement between the observed cases and the simulated outbreaks using the optimal parameters. The effective reproduction number also reflects the effects of the interventions at Harvard and OSU in way that’s consistent with our initial modeling assumptions.

#### 3.1.1 Maximum Likelihood Estimates for Harvard

The results are shown in Table 2. Notably, the baseline removal rate *γ*_*H*_ is quite high, indicating that the initial diagnosis protocol was quite effective at identifying and removing infected students from the population, but it was further increased after day 61. The reporting rate *ρ*_*H*_ is also remarkably high, which suggests that HUHS was able to identify most of the cases circulating at Harvard.

**Table 2:** List of parameters in the Harvard model that were obtained by MLE or calculated using the estimated parameters.

| *Symbol* | *Description* | *Point estimate* | *95% CI* | *Units* | *Source* |
| --- | --- | --- | --- | --- | --- |
| $\beta_H$ | Baseline transmission rate | 1.39 | (1.02, 2.20) | day<sup>-1</sup> | MLE |
| $\gamma_H$ | Baseline removal rate | 0.85 | (0.78, 0.99) | day<sup>-1</sup> | MLE |
| $p$ | Decrease in infection due to vacation | 0.11 | (0.00, 0.47) | | MLE |
| $q$ | Increase in removal rate | 2.8 | (1.39, 6.00) | — | MLE |
| $\rho_H$ | Proportion of infections reported | 0.97 | (0.87, 0.99) | — | MLE |
| $\psi_H$ | Overdispersion parameter | 0.54 | (0.36, 0.72) | — | MLE |
| $R_E(t)$ | Effective reproduction number | 1.63 normal term<br>0.18 during vacation<br>0.58 after intervention | — | — | Calculated as $\frac{\beta(t)}{\gamma(t)}$ |

We ran 200 stochastic simulations of Harvard’s outbreak using the parameter values from Table 2. Figure 3A shows the range of values across simulations, and they appear consistent with the observed data. Shortly after day 61 (the time of the primary intervention), we see a decrease in the number of cases. The variability in the simulations can partly be attributed to the randomness in the stochastic model as well as the over-dispersion parameter. Variability can also be explained by the MLE of the basic reproduction number being below 2, which together with the stochasticity built into the simulations, can result in absence of outbreak.

**Figure 3:**
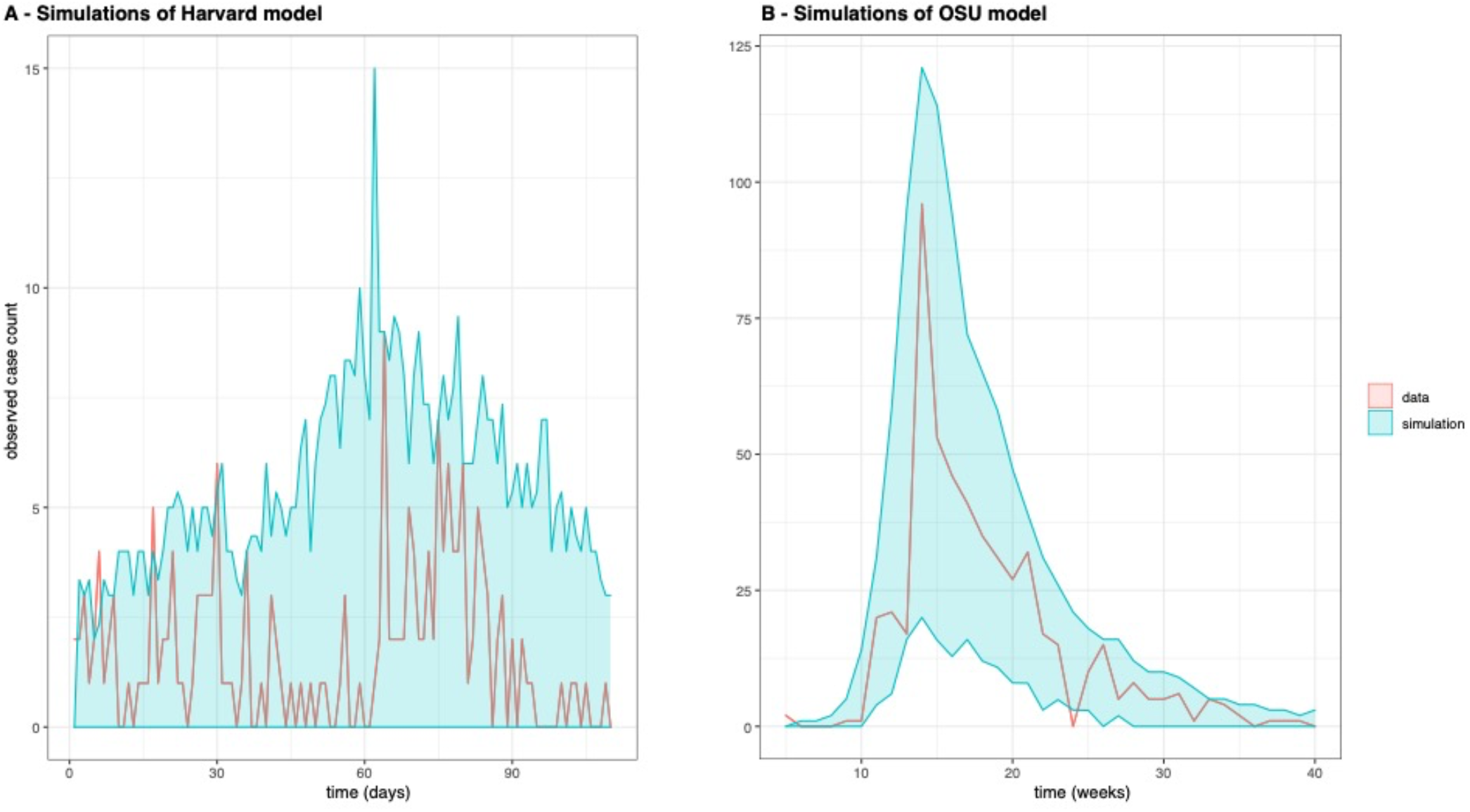
These plots show the observed case count data (red line) and the range of simulated case count values at each time point between the bottom 5% and top 95% percentiles (blue shaded area) from 200 simulation runs using the Harvard (A) and OSU (B) models evaluated at the maximum likelihood estimates of the parameters.

#### 3.1.2 Maximum Likelihood Estimates for OSU

The MLEs of the parameters for the OSU model, as well as derived quantities, are shown in Table Here we can see an initial reproductive number of almost 6, much higher than Harvard’s. However, it eventually becomes lower than 1, which supports our modeling assumptions of an awareness campaign from OSU, perhaps helped by news reporting about the outbreak, that lead to effective self-isolation of individuals.

As with Harvard, we run stochastic simulations of OSU’s outbreak using the parameter values from Table 3. The simulated outbreaks are shown in Figure 3B, and they follow the real data remarkably well. However, the proportion of infections reported, *ρ*_0_, is very low at 3%. This would imply that the true number of cases in the OSU outbreak was 30 times larger than observed, placing the total count at around 15,000.

**Table 3:** List of parameters in the OSU model that obtained by MLE or calculated using the estimated parameters.

| *Symbol* | *Description* | *Point estimate* | *95% CI* | *Units* | *Source* |
| --- | --- | --- | --- | --- | --- |
| $\beta_O$ | Transmission rate constant | 1.19 | (0.92, 1.43) | day<sup>-1</sup> | MLE |
| $w$ | Decrease in infection due to self-isolation | 0.16 | (0.09, 1.17) | — | MLE |
| $\rho_O$ | Proportion of infections reported | 0.03 | (0.03, 0.08) | — | MLE |
| $\psi_O$ | Overdispersion parameter | 0.38 | (0.22, 0.65) | — | MLE |
| $R_E(t)$ | Effective reproduction number | 5.95 initial<br>0.95 after advisory | | — | Calculated as $\frac{\beta(t)}{\gamma(t)}$ |

### 3.2 Earlier intervention decreases outbreak size at Harvard and OSU

The results from the intervention analysis for Harvard and OSU is depicted in Figure 4. By the final day of the Harvard outbreak (day 130), the simulations without the intervention on day 61 yielded outbreak sizes that were up to four times the size of the actual outbreak (Figure 4A). These results also indicate that the outbreak would have lasted much longer, if not for these vigilance-increasing strategies. By varying the day of the intervention from 1 to 61, we also obtained a linear regression between day of intervention and reduction of the outbreak (Figure 4C). The fitness of the regression is very high (R^2^=0.96, P<10^−9^), and quick inspection of the plot reveals that if the new diagnosis protocol had been implemented within the first 10 days of the outbreak, then no more than 50 students would have been infected in total at Harvard.

**Figure 4:**
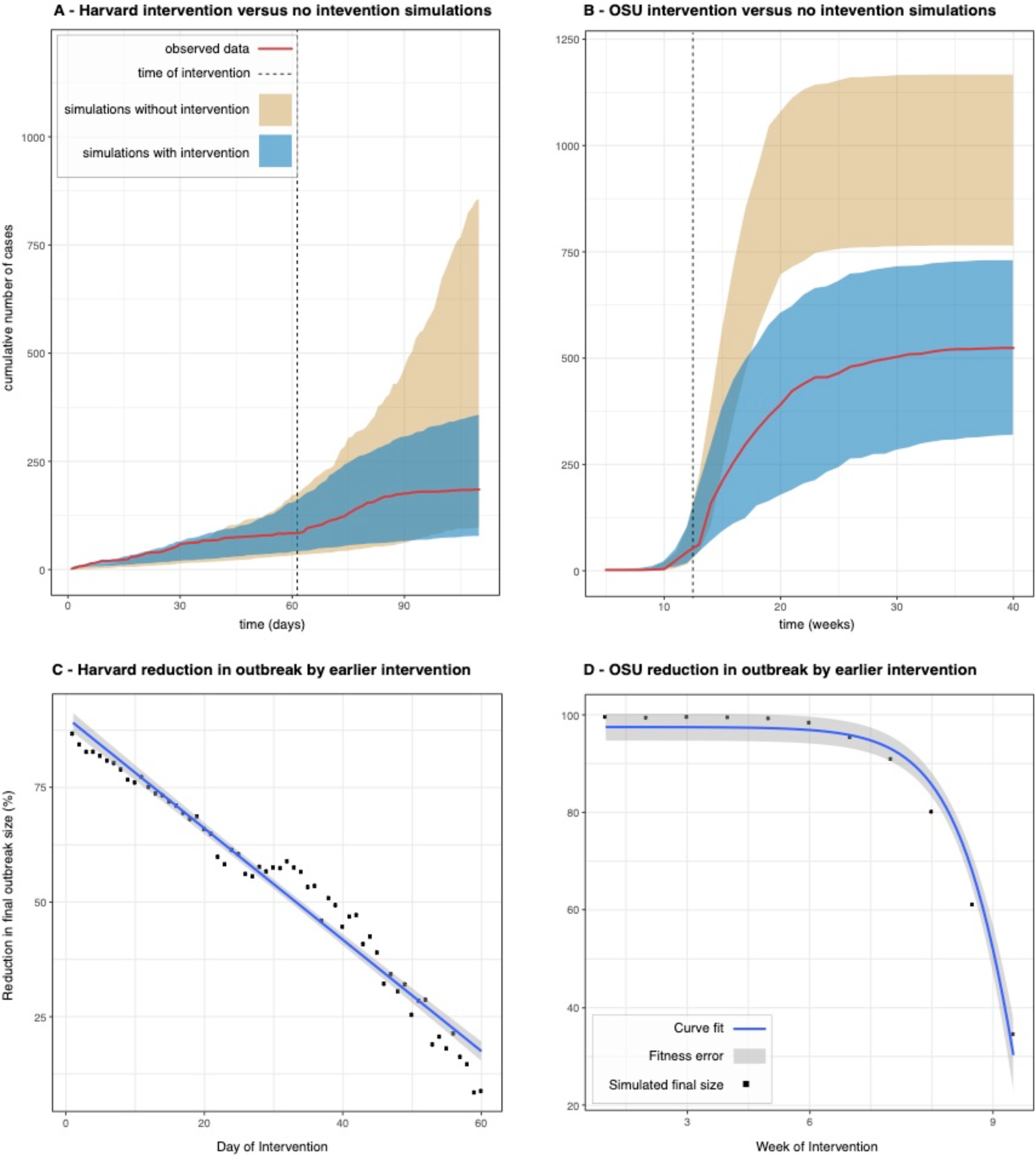
Panels A and B show the comparison of the cumulative number of cases over time for the observed Harvard and OSU data and the range of cases (95% percentile of the runs) in simulations with and without interventions, with dotted lines representing the timing of the interventions in each school (panels A and B). In panels C and D, the plots show the percentage we expect outbreak size to decrease by if the date of intervention had been moved up. There is a significant linear relationship between the time and percentage reduction in the case of Harvard, as well as a significant relationship after doing a sigmoid transformation of the time variable in the case of OSU.

For OSU we observe similar trends. Lack of intervention on week 12 could have resulted in an outbreak twice as large (Figure 4B). The outbreak size as a function of the intervention week also shows a strong dependency, but in this case non-linear and best fit with a sigmoid function of the form 1/(1+e^week-12^). Using this transformation, the fit is also very high (R^2^=0.63, P<0.005), and we can conclude that intervening earlier would have had a major effect as well: if the awareness campaigns prompting students to self-isolate had started around week 5 or 6 (rather than week 12), then it appears likely that the outbreak could have been completely eradicated.

## 4. Discussion

### 4.1 Parameter interpretation

The MLEs give us insight into characteristics of the mumps outbreaks at Harvard University in 2016 and Ohio State University in 2014, as measured by their effective reproduction numbers *R*_*E*_, intervention parameters *q* and *w*, rates of removal *γ*, reporting rates *ρ*, and overdispersion parameters *ψ*. At Harvard, *R*_*E*_ during normal class term was 1.63, which indicates that the outbreak was growing, even though testing and isolation by HUHS resulted in a baseline removal time of only 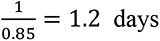. This points to the effectiveness of the quarantine system implemented by HUHS. However, a small fraction of false negative cases still managed to escape quarantine and keep the virus under circulation, as indicated by the reproduction number being higher than 1. The reproduction number goes below 1 during the spring break, which is reasonable given that most students are away due to the residential nature of the Harvard campus. However, transmission resumes after the break. It is only after the implementation of the new diagnosis protocol on day 61, which required isolation if clinical symptoms were present, that had a dramatic effect on the detection and isolation of positive cases, effectively taking the removal time to less than 1 day and the reproductive number below 0.6. Thanks to this key intervention, it was possible to end the outbreak before the beginning of the summer recess.

The estimate of *ρ* is 0.96, which implies the reporting rate at Harvard was remarkable. Reasons include the email awareness campaign, a community network – from resident deans to athletic coaches – reporting students and employees who seemed at-risk, and more aggressive diagnoses, particularly towards the end of the outbreak. The estimate for *ψ* is 0.54, suggesting that the actual data has more variability than expected under the assumed distribution. If *ψ* had been approximately 0, the variance in our measurement model would have simplified to the variance for a binomial distribution. However, because the 95% confidence interval is (0.22, 0.65) and thus does not include 0, we justify the modelling decision of representing the number of cases as an over-dispersed binomial. Demographic and environmental stochasticity (e.g.: a student during midterm season may be less likely to report symptoms), as well as the interventions themselves (e.g.: reporting may increase temporarily after an awareness email) can result in over-dispersion in the number of reported cases.

In the case of OSU, we obtain a much higher reproduction number at the beginning of the outbreak, near 6, and a very low reporting rate of 3%. Before discussing these results any further, it is important to keep in mind that we extrapolated OSU cases from state-level reports by the CDC. Furthermore, we did not have direct access to information about the containment interventions adopted by the school, as we did for Harvard, so we were only able to make educated guesses about those possible interventions based on information we found on the web. Within our OSU model, we can conclude that self-isolation of students motivated by the advisories posted by OSU had the intended effect of stopping the outbreak. The effective reproduction number dips below 1 after March, which is when the awareness campaign appeared to have started, and when the outbreak gained local and national prominence due to news reporting. A significant issue with the OSU model is the very low reporting rate of 3% derived from the MLE calculation. This rate implies that the true number of mumps cases during the OSU outbreak should have been approximately 30 times larger than observed. It follows that the total number of cases could have reached 15,000 individuals, which is internally consistent in the model given that the number of susceptible within the school’s student population is over 30,000. However, such a large case count is unlikely, as it doubles the highest number of yearly mumps cases reported in the US in the last 20 years (38), which was 6,369 in 2016. There is little data on the percentage of asymptomatic mumps infections that would result in non-reporting, but the available evidence points to 15%– 30% (39, 40). Seroprevalence studies from the pre-vaccination era indicate a reporting rate of around 4% (41), which is strikingly similar to the MLE estimate for OSU, but it is hard to imagine such level underreporting in 2014 and in the midst of an active outbreak. Therefore, we are not confident on this parameter estimate, even though the 95% confidence interval is very narrow at (3%, 8%). Our interpretation of this situation is that the modeling approximation of a closed SEIR compartments is probably less accurate for OSU given its non-residential nature: students there have more opportunity to interact with individuals outside of their school, resulting both in additional transmissions that are not captured by our model, and also in a “buffering” effect due to a largely immune population (outside the college age).

### 4.2 Effect of strict isolation policy

Arguably the most critical intervention by HUHS was the isolation requirement for confirmed and probable mumps cases. By taking the Harvard model on its own, we see that the infectious period was already quite low at 1.2 days, even before the update in testing protocols. This conclude that the isolation policy led to a smaller average infectious period for Harvard patients. The MLEs for Harvard and OSU are different for several parameters, most notably basic reproduction number, reporting rate, and rate of transition from the infectious to removed class. Firstly, OSU’s basic reproduction number is over four times that of Harvard. Harvard’s isolation policy best explains this difference because it physically prevented infectious persons from causing multiple secondary infections, thus suppressing the growth of the outbreak. But as pointed out before, the extremely low reporting rate inferred from the OSU model makes us less confident in it, so any comparison between the Harvard and OSU outbreaks based on the MLE parameters should be taken with caution.

### 4.3 Implications of intervention analysis

With the benefit of our intervention analysis, we conclude that aggressive diagnoses decreased the size of the Harvard outbreak by approximately three-fourths. Furthermore, for every day of intervention delay, we estimate that the outbreak size would have increased by 1.6 percentage points, extrapolating the regression line in Figure 4C. Likewise, self-isolation prompted by health advisories posted by the university reduced the size of the OSU outbreak by half. Given the non-linear dependency between change in outbreak size and timing of intervention (Figure 4D), the increase would have been even larger in that outbreak. Interestingly, this dependency also implies that self-isolation in the first weeks of the outbreak can be enough to completely stop spread.

Clearly, a limitation of this analysis is the assumption that everything remains the same while changing the time of the intervention under consideration. In reality, other factors might come into play if the outbreak becomes larger or smaller, which in turn could affect the dynamics of the outbreak as well as the interventions themselves. However, this analysis still provides a useful hypothetical quantification of the effect of accelerating or delaying interventions designed to contain the spread of an outbreak and here, as expected, the sooner the interventions are introduced, the better the outcomes in terms of outbreak size. Of course, existing constraints in the school’s health system could impede fast interventions. In such situations, our method can be useful to perform a cost-benefit analysis of how late an intervention could be made to still have a significant reduction in the health burden caused by the disease.

### 4.4 Conclusions

We constructed and parametrized a POMP model for the transmission of mumps on college campuses. The POMP model is a computationally efficient approach suitable to small populations that accounts for the noisiness and incompleteness of case data. Moreover, it incorporates parameters that measures the effect of interventions implemented after a given point in time. Given the worldwide crisis caused by the COVID-19 pandemic, such models can be useful to quickly evaluate interventions designed to contain the spread of SARS-CoV-2 once schools reopen in the U.S. and around the world.

We compared an outbreak at Harvard University, with its various intervention strategies, to another university outbreak of comparable reported cases at OSU. Importantly, while most literature today focuses on mumps prevention – such as administering third MMR doses to college-age students – this paper provides quantitative backing for more immediate and less costly approaches to mitigating the spread of mumps and other infectious diseases, most notably COVID-19. Even with widespread availability of vaccines, outbreaks of highly transmissible diseases are still a reality, as mumps exemplifies very clearly. In particular, requiring strict isolation if any symptoms of the disease are presented would significantly reduce transmission and ultimately the size of the outbreak. Effective awareness campaigns that lead to self-isolation of infected individuals with mild symptoms can also have a significant effect in containing the spread of disease and limiting the risk for vulnerable populations.

### 4.4 Limitations

Some of our conclusions are likely affected by confounding factors that we cannot control for in this analysis. For example, the outbreak at Harvard started to subside in late April, not long before students finish the semester and leave campus, which would decrease the number of potential infections. The most promising method to determine the exact effect of isolation strategies is through a randomized control trial. Regarding the differences between OSU and Harvard parameters, we must be cautious in taking the OSU estimates at face value given the inconsistency in the reporting rate, which may be indicating a more fundamental limitation of model to represent the OSU outbreak. In addition to that, given that the OSU data consists of weekly reports rather than daily reports of cases, we should expect the estimates for the parameters to be less accurate. Furthermore, the cases are not solely linked to the university. Numerous cases in the data occurred in the greater Columbus area, suggesting that the parameter estimates do not only account for the dynamics of mumps on campus. Lastly, major differences in housing and campus characteristics could have also contributed to differences between the two schools; for instance, OSU’s population size is three times that of Harvard, and OSU has larger dorms than Harvard’s houses. Interventions used at Harvard simply may not have worked as well at OSU. We were fortunate to have direct access to school administrators who were involved in the response to the 2016 outbreak to discuss HUHS interventions in detail, but we were not able to get the same level of detail for OSU’s interventions, as discussed in the main text. More broadly, lack of publicly available datasets, with the exception of CDC reports on OSU’s outbreak, is a serious impediment to perform these analyses. Therefore, it will be essential that universities across the US and globe actively share data for comparative analysis, to identify the best intervention strategies to protect college campuses from outbreaks, especially in the post-COVID-19 world.

## Data Availability

Data is freely available online

https://github.com/broadinstitute/mumps-pomp-models

## Source Code

Available at https://github.com/colabobio/mumps-pomp-models

## Author’s Contributions

MS participated in the design of the study, carried out the data analysis, developed the epidemiological models, generated the conclusions, and drafted the manuscript; GF developed the epidemiological models, and generated the conclusions; AC conceived of the study, participated in the design of the study, coordinated the study, and helped draft the manuscript; SF and PJB provided data on the HUHS interventions and reviewed the final draft of the manuscript; PCS overviewed the study and reviewed the final draft of the manuscript. All authors gave final approval for publication and agree to be held accountable for the work performed therein.

## Acknowledgements

The authors would like to thank Jonathan Grad and Joseph Lewnard for providing feedback on the study design, Hayden Metsky for reviewing the manuscript, members of MDPH for providing access to the Harvard data, and Bridget Chak and Shirlee Wohl for guiding in the interpretation of the data.

## Ethics

Usage of Harvard University data for development of the SEIR model was approved by the Massachusetts Department of Public Health (MDPH) through protocol 906066. Harvard University Faculty of Arts and Sciences and the Broad Institute ceded review of secondary analysis to the MDPH IRB through institutional authorization agreements. The MDPH IRB waived informed consent given this research met the requirements pursuant to 45 CFR 46.116 (d). Data from Ohio State University was obtained from the CDC’s Morbidity and Mortality Weekly Report 2014. The Broad Institute has determined usage of this data constitutes non-human subjects research.

